# Significant rise of *Chlamydia pneumoniae* diagnoses at Marseille University Hospitals, 2024, France

**DOI:** 10.1101/2024.12.11.24318723

**Authors:** Sophie Edouard, Rayane Attamna, Matthieu Million, Céline Boschi, Jeremy Delerce, Aurélia Caputo, Didier Stoupan, Seydina Diene, Idir Kacel, Claudia Andrieu, Anthony Levasseur, Hervé Chaudet, Jean-Marc Rolain, Lucile Lesage, Aurélie Morand, Pierre-Edouard Fournier, Jean-Christophe Lagier, Florence Fenollar, Bernard La Scola, Philippe Colson

## Abstract

We report a 19-fold increase in 2024 of qPCR diagnoses of *Chlamydia pneumoniae* infections in Marseille, Southern France, with 37 cases versus 10 between 2018-2023. These mostly affected children, and young adults. We obtained four *C. pneumoniae* genomes, all of serotype ST16, suggesting an epidemic circulation in our geographical area.

## MAIN TEXT

Respiratory infections are a major cause of morbidity and mortality worldwide regardless of the geographic area. The SARS-CoV-2 pandemic profoundly changed the epidemiology of respiratory infections. This was the case in France, including in Marseille, Southeastern France, with a quasi-absence of influenza diagnoses during winter 2020-2021 or a shift of several months of the epidemic period of respiratory syncytial virus (RSV) in 2021 (1,2). In addition, such changes in the incidence of infections were also observed for microorganisms including some involved in respiratory diseases (3).

Recently, dramatic increases in the incidence of respiratory microbial infections, due to *Mycoplasma pneumoniae* (4) then *Bordetella pertussis* (5) have been observed in France in 2023-2024. *C. pneumoniae* is another bacterium associated with acute respiratory infections and causing from upper respiratory tract mild disease to pneumonia. It is implicated in <1.5% of cases of community-acquired respiratory infections and particularly affects children (6). It usually causes asymptomatic or mild infections but severe cases, asthma exacerbation and chronic respiratory illness can occur (7). Its diagnosis relies mainly on real-time PCR (qPCR). Here we describe a significant recent increase in qPCR diagnoses of *C. pneumoniae* respiratory infections, and the epidemiological, microbiological, and genomic characteristics of these infections.

### The study

We detected in October 2024 an abnormally high number of *C. pneumoniae* diagnoses. This rise triggered the implementation of an automatic daily report transmissible to hospital biologists and clinicians through a basic mailing list and a configurable subscription. We retrospectively analyzed *C. pneumoniae* qPCR results performed in the clinical microbiology diagnosis laboratory of university/public hospitals of Marseille, Southern France, on respiratory samples collected from patients between 01-Jan-2018 and 30-Oct-2024. qPCR was either part from a multiplex syndromic panel (Biofire FilmArray Respiratory panel 2, bioMérieux, Marcy-l’Etoile, France) used at point-of-care laboratories or was an in-house simplex assay targeting the *omp2* gene (8) (see more detailed procedures in Appendix Methods). Co-infection with other respiratory pathogens was assessed using results of the Biofire assay or of the Fast Track Diagnosis Respiratory pathogens 21 assay (Fast Track Diagnosis, Luxembourg). Next-generation sequencing (NGS) was performed on residues of five samples with the lowest known qPCR cycle threshold values (Ct) using Oxford Nanopore (Oxford Nanopore Technologies, Oxford, UK) and Illumina technologies (Illumina Inc., San Diego, California, USA), on GridION, and MiSeq and NovaSeq 6000 instruments, respectively. Bioinformatic analyses of NGS reads and phylogenomic analyses were performed as detailed in Appendix Methods. OpenEpi version 3.01 (https://www.openepi.com/) was used for statistical analyses (*p*-values ≤ 0.05 were considered significant).

Overall, 35,344 specimens collected from patients between 01-Jan-2018 and 30-Oct-2024 had been tested, including 30,848 upper and 4,496 lower respiratory samples. Among them, 19,504 samples were collected in men (55%). Median age of patients was 5-year-old (range, 0-107). Overall incidence of *C. pneumoniae* qPCR-positivity was 0.13% (47/35,344 being positive). During 2018-2022, the number of samples tested for *C. pneumoniae* by qPCR regularly increased, with a peak at 11,754 samples in 2022 (Figure 1a). However, only five samples were positive during this time lapse. In 2023, Biofire assay use was restricted, which was associated with a drop of the total number of *C. pneumoniae* qPCR tests to 4,294 and 5,795 samples in 2023 and 2024, respectively. Nonetheless, in 2024, 37 patients (0.64%) were diagnosed compared to 5 (0.12%) in 2023 (p<0.0001). Five cases occurred between January-June, then six in July-August, 15 in September, and 6 in October 2024 (Figure 1b).

**Figure 1.**
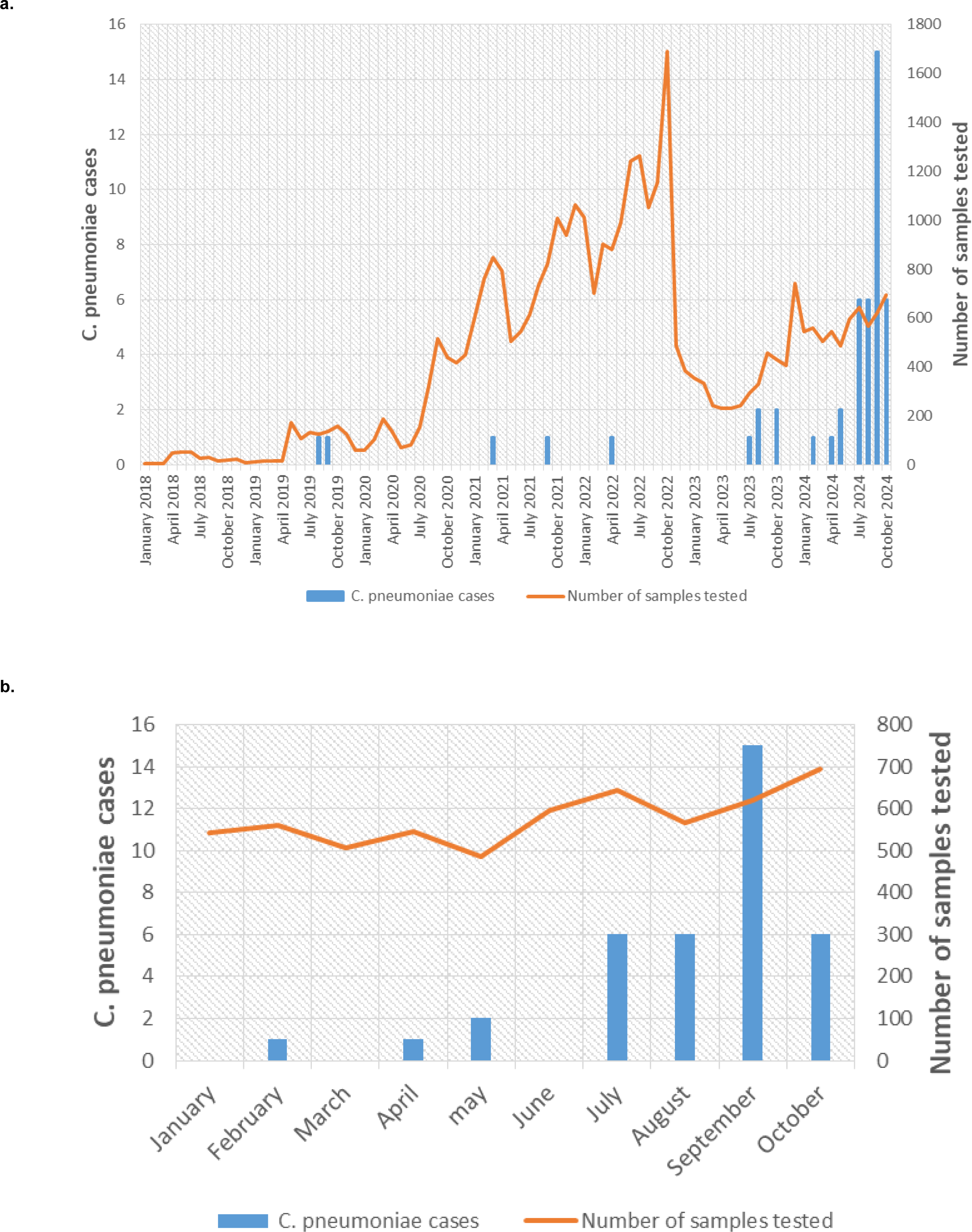
Number of respiratory samples tested and positive by a *Chlamydia pneumonia* qPCR assay between 2018 and 2024 (a) and during Year 2024 (b)

The 37 specimens positive in 2024 included 22 nasopharyngeal swabs, 11 nasopharyngeal aspirates and 4 sputums. Twenty-eight (76%) were initially diagnosed positive using the Biofire assay. None of the 4,496 low respiratory samples tested positive. Among the 37 patients, 19 (51%) were male; median age was 10-year-old (range, 0-44). Twenty-seven (73%) patients were children <18-year-old and 10 (27%) were young adults. Median age was 8 years (range, 2 months-15 years) in children and 34 (25-44) in adults. Children <1 year, 6-10 years and 11-15 years were the age groups the most represented among *C. pneumoniae* cases, comprising each between 19-22% of these cases (Table 1). No patient >50 years was diagnosed in 2024. Thirteen patients were admitted in hospitalization, including six for other reasons than severity of respiratory symptoms; no case was admitted in intensive care unit or died. Fourteen (38%) of the 37 patients were coinfected with ≥1 other respiratory pathogen, including rhinovirus (n=9), SARS-CoV-2 (n=2), parainfluenza virus (n=1), coronavirus-NL63 (n=1), adenovirus (n=1), coronavirus-HKU1 (n=1), and *Bordetella pertussis* (n=1) (Figure 2).

**Figure 2.**
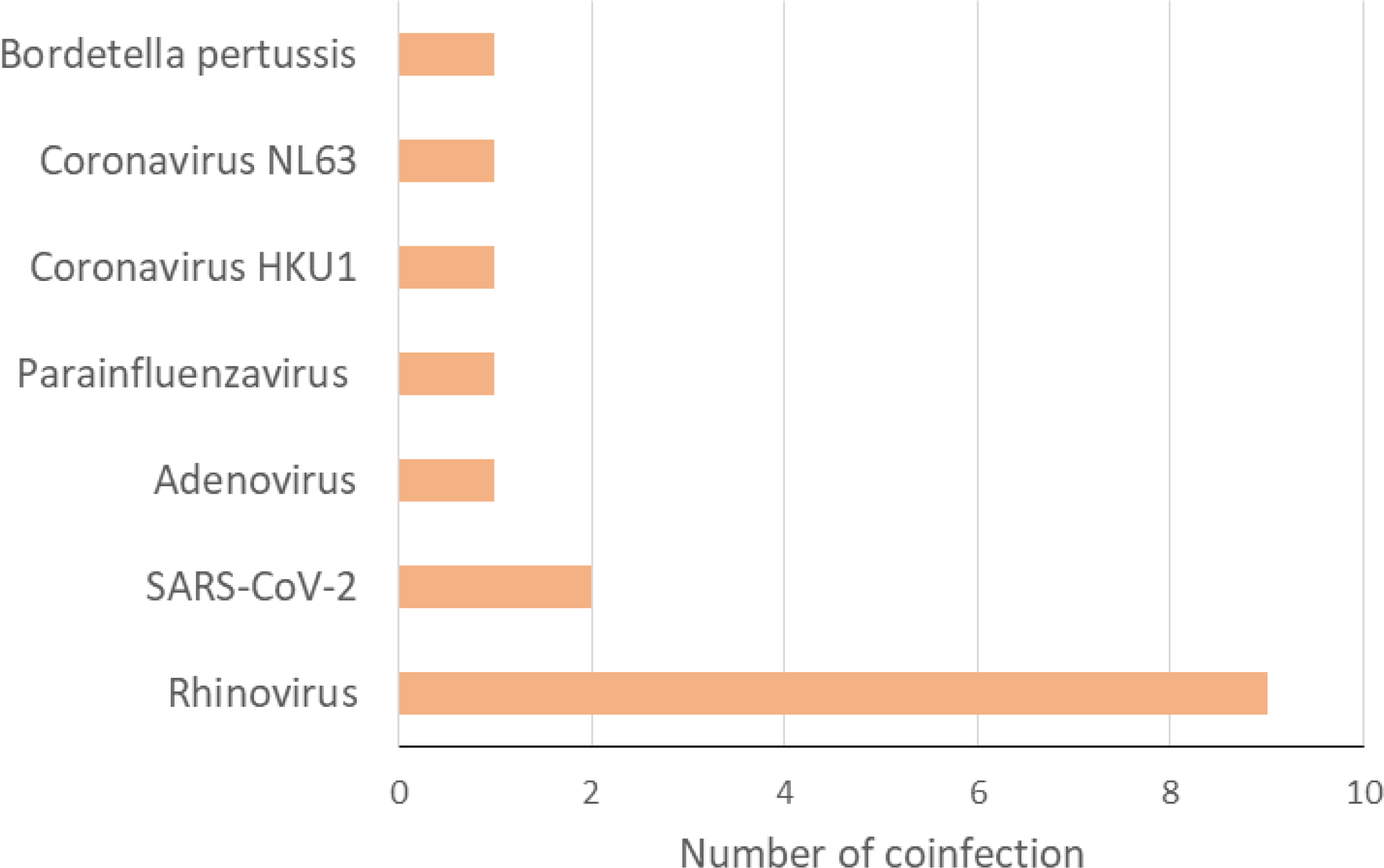
Number of coinfections with other infectious agents for respiratory samples positive by a *Chlamydia pneumoniae* qPCR assay in 2024

**Table 1.** Demographics characteristics and coinfections of the cases of *C. pneumoniae* infection diagnosed by qPCR in our center in 2024.

| Characteristics | n (%) |
| --- | --- |
| **Incidence of *C. pneumoniae* cases** | 37/5795 (0.6) |
| ***C. pneumoniae* cases** |  |
| Male | 19/37 (51.4) |
| Female | 18/37 (48.6) |
| Median age (years) | 10 |
| Age distribution (years) |  |
| <1 | 7/37 (18.9) |
| 1-5 | 4/37 (10.8) |
| 6-10 | 8/37 (21.6) |
| 11-15 | 8/37 (21.6) |
| 16-20 | 0/37 (0.0) |
| 21-35 | 6/37 (16.2) |
| 36-50 | 4/37 (10.8) |
| > 50 | 0/37 (0.0) |
| Number of coinfection | 14/37 (37.8) |
| **Incidence of *C. pneumoniae* infections by age groups** |  |
| <1 | 7/11,215 (0.1) |
| 1-5 | 4/7,074 (0.1) |
| 6-10 | 8/1,669 (0.5) |
| 11-15 | 8/1,075 (0.7) |
| 16-20 | 0/606 (0.0) |
| 21-35 | 6/1,943(0.3) |
| 36-50 | 4/2,063 (0.2) |
| > 50 | 0/9,698 (0.0) |

Two *C. pneumoniae* whole circular genomes (1,231,868 and 1,232,097 base pair (bp)-longs, in one scaffold) (GenBank Accession no. CP172581 and CP173416), two additional near full-length genomes in 11 or 12 scaffolds (1,229,923 and 1,231,291 bp-longs) and one partial genome in 65 scaffold (306,688 bp) were obtained by NGS directly from respiratory samples (Appendices Results and Table 1). All four (near) full-length genomes were classified as ST16 based on MultiLocus Sequencing Typing (Appendices Methods, Table 2), suggesting the circulation of a single serotype in 2024. Only 19 other *C. pneumoniae* genomes were available from GenBank (https://www.ncbi.nlm.nih.gov/genbank/), six being classified as ST16. Phylogenomics indicated that the two full-length genomes obtained here were those the most closely related with each other (Appendices Figures 1a,b).

### Conclusion

Global systematic surveillance of diagnoses at our institution allowed detecting a *C. pneumoniae* ST16 outbreak in 2024 in Marseille, France, which peaked in September. As previously described for *M. pneumoniae* then *B. pertussis* (4,5), we observed that *C. pneumoniae* qPCR positivity rate raised to a level never seen before in our center. Most cases affected children and no severe case was reported. As commonly observed in other respiratory infections, we found a high rate of coinfection of *C. pneumoniae* with other pathogens (2,4,6).

A resurgence of several respiratory pathogens was observed following the global attenuation of the SARS-CoV-2 pandemic (1,3,4-6). Increase of positive *C. pneumoniae* qPCR cases after the SARS-CoV-2 pandemic was previously observed in 2023 in Switzerland (6); no genome sequencing was reported. *C. pneumoniae* epidemics were also described before the SARS-CoV-2 pandemic, such as in a prison in Texas in 2014 (9) and among children hospitalized with community-acquired pneumonia in Korea in 2016 (10).

The number of *C. pneumoniae* cases is likely underestimated in our study, particularly because patients with mild symptoms are not systematically tested and *C. pneumoniae* is not part of the respiratory syndromic qPCR panel used at our core laboratory. Indeed, samples from ≈2,000 patients presenting respiratory diseases and requiring routine diagnostic testing between July-October 2024 were not specifically tested for *C. pneumoniae*. Moreover, the amount of asymptomatic cases that may play a role in the epidemic is unknown. Here, we did not consider *C. pneumoniae* serological results due to the lower sensitivity and specificity of serology compared to qPCR and to serum unavailability for most case-patients.

Unexpectedly, the number of other *C. pneumoniae* genomes released in GenBank was very few, considering the frequency and pathogenicity of this bacterium, and the sharp interest of microbial genomic surveillance for epidemiology and clinics. Among the 19 previously available *C. pneumoniae* genomes, the most frequent genotype was ST17 (n=11) followed by ST16 (n=6).

Overall, present data warrants surveying closely diagnoses of *C. pneumonia*e infections at the local and country scales; and implementing genomic surveillance and characterizing drug resistance for diagnosed cases.

### First author’s short biographical sketch

Dr. Sophie Edouard is a medical bacteriologist at the Microbiological Laboratory at IHU Méditerranée Infection in Marseille, France. Her research interests focus on infectious diseases and microbiology including intracellular bacteria and emerging pathogens.

## Acknowledgements

We are very grateful to the technical team of the next-generation sequencing platform of the IHU Méditerranée Infection.

## Data availability

*Chlamydia pneumoniae* genomes in one scaffold analyzed here have been submitted to the GenBank sequence database (https://www.ncbi.nlm.nih.gov/genbank/) (11) (GenBank Accession no: CP172581 and CP173416).

## Author contributions

Conceived and designed the experiments: SE, PC. Contributed materials, analysis tools: All authors. Analyzed the data: SE, JD, AC, SD, PC. Writing—original draft preparation: SE, PC. Writing—review and editing: All authors. All authors have read and agreed to the published version of the manuscript.

## Ethics

The present study involves data that have been registered on the Health Data Access Portal of Marseille university and public hospitals (Assistance Publique-Hôpitaux de Marseille (AP-HM)) under No. PADS24-337, and was approved by the AP-HM Ethics and Scientific Committee.

## APPENDICES

### APPENDIX, SUPPLEMENTARY METHODS

qPCR was either part from a multiplex syndromic panel (Biofire FilmArray Respiratory panel 2; bioMérieux, Marcy-l’Etoile, France) used at point-of-care laboratories operating 24/24 7/7 for urgent requests, or was an in-house simplex qPCR assay targeting the *omp2* gene used at the core laboratory operating only on working days (1). Co-infections with other respiratory pathogens were assessed using results of the Biofire FilmArray assay, or of the Fast Track Diagnosis Respiratory pathogens 21 assay (Fast Track Diagnosis, Luxembourg) performed at the core laboratory on working days.

Next-generation sequencing (NGS) was performed at the IHU Méditerranée Infection clinical microbiology and virology laboratory on residues of samples with the lowest qPCR cycle threshold values (Ct) using (i) Oxford Nanopore technology with the Ligation Sequencing Kit (SQK-LSK109) and a SpotON flow cell Mk I, R9.4.1 on a GridION instrument (Oxford Nanopore Technologies, Oxford, UK); and (ii) Illumina technology using the classic Illumina loading procedure in XP mode on a MiSeq and a NovaSeq 6000 instruments (Illumina Inc., San Diego, California, USA), with a reading of 2x250 and 2x150, respectively.

Nanopore reads were filtered with filtlong (v0.2.1) (https://github.com/rrwick/Filtlong) while Illumina reads were filtered using fastp (v0.23.4) (2). Only the minimum read length parameter was modified, to 1,000 bp for the Nanopore runs, and to 100 base pairs (bp) for the Illumina runs. Cleaned reads were then classified with Kraken2 (v2.0.9-beta) (https://github.com/DerrickWood/kraken2) (3) using the “standard” database from 2024-05-06, with a confidence threshold of 0.02. Hits corresponding to taxon 83558 (including its offspring) were extracted using KrakenTools (v1.2) (https://github.com/jenniferlu717/KrakenTools) (4). Reads corresponding to *Chlamydia* spp. were assembled using Unicycler (v0.5.1) (https://github.com/rrwick/Unicycler) (5), Raven (https://github.com/lbcb-sci/raven) (6), and Polypolish (https://github.com/rrwick/Polypolish) (7). The resulting genomes were annotated with Bakta (v1.9.4) (https://github.com/oschwengers/bakta) (8) using the “5.1 full” database, and Roary (https://sanger-pathogens.github.io/Roary/) (9). Pangenomic analysis was performed with PIRATE (v1.0.5) (https://github.com/SionBayliss/PIRATE) (10) using the -a option to generate the core alignment. The core genome phylogeny was inferred using IQ-TREE2 (http://www.iqtree.org/) (11) with the -m GTR+R option. The pangenome was visualized with Phandango (https://github.com/jameshadfield/phandango) (12), and the phylogenetic tree, including bootstrap values, was annotated with iTOL (https://itol.embl.de/) (13). The MultiLocus Sequencing Typing (MLST) analysis was performed using the PubMLST web application (https://pubmlst.org/software/bigsdb) (14) and its database for *Chlamydia* spp. *C. pneumoniae* genomes were automatically annotated by the NCBI prokaryotic genome annotation pipeline (15). Resistance genes were searched using the Arg-Annot tool (16).

### APPENDIX, SUPPLEMENTARY RESULTS

The abnormally high number of *C. pneumoniae* diagnoses was detected in October 2024 in our institute. This was discussed during our daily diagnoses report meetings that involve all medical biologists and medical biology residents of the clinical microbiology and virology laboratory of Marseille university hospitals at IHU Méditerranée Infection, and during our weekly meetings on epidemiological surveillance of infections based on the data of this laboratory, to which attend medical biologists and clinicians from our infectious diseases institute (17). Specific daily automated surveillance of new diagnoses of *C. pneumoniae* that included the localization in clinical wards of hospitalized cases was newly implemented in October 2024, and transmitted each day to medical biologists and clinicians from Marseille university hospitals, and the rise in the *C. pneumoniae* diagnoses was reported to the regional health agency.

Cycle threshold values (Ct) of our in-house qPCR were available for 28 of the 37 *C. pneumoniae*-positive samples and ranged between 18 and 37. The overall incidence of *C. pneumoniae* cases was higher in children between 11-15 years (8/1,075, 0.7%) and in children between 6-10 years (8/1,669, 0.5%), compared to the other age groups (Main text Table 1). Children <1 year were more frequently coinfected (86%) with ≥1 other respiratory pathogen compared to the other age groups (Main text Table 1).

*C. pneumoniae* genomes were obtained by NGS performed directly from respiratory samples with Ct lower than or or equal to 22 (range, 18-22). The two whole genomes in a single scaffold were obtained from samples with a qPCR Ct of 21. They harbored between 1,098 and 1,101 genes. Genes encoding hypothetical proteins and DUF-domain containing proteins were those the most frequent, in between 210 and 217 and 64 and 65 cases, respectively. All five genomes obtained here belonged to ST16 based on MultiLocus Sequencing Typing (Appendix Table 2), suggesting the circulation of a single serotype during this 2024 outbreak. Only 19 other *C. pneumoniae* were available Phylogenomic analyses based on mutations in core genes and conducted for the four (near) full-length *C. pneumoniae* genomes (in between 1-12 scaffolds) obtained here and the 19 *C. pneumoniae* genomes previously released in the GenBank NCBI nucleotide sequence database (https://www.ncbi.nlm.nih.gov/genbank/) (18) indicated that the four (near) full-length genomes obtained in our laboratory comprised a cluster, being those the most closely related with each other (Appendices Figures 1a,b). Only 16 mutations differentiated the two whole *C. pneumoniae* genomes in a single scaffold obtained here based on the alignment of core genes. Six other genomes from GenBank were classified as of ST16, the most frequent serotype being ST17 (n=11). Resistance gene search revealed the presence of a gene encoding an UDP-N-acetylglucosamine 1-carboxyvinyltransferase (murA) potentially associated with fosfomycine resistance, which was only described to our knowledge in *Chlamydia trachomatis* genomes among *Chlamydia* spp. (19), and was shared by the other *C. pneumoniae* genomes. No fluoroquinolone, macrolide or tetracycline resistance genes were found.

### APPENDIX, SUPPLEMENTARY FIGURES

**Appendix, Figure 1.**
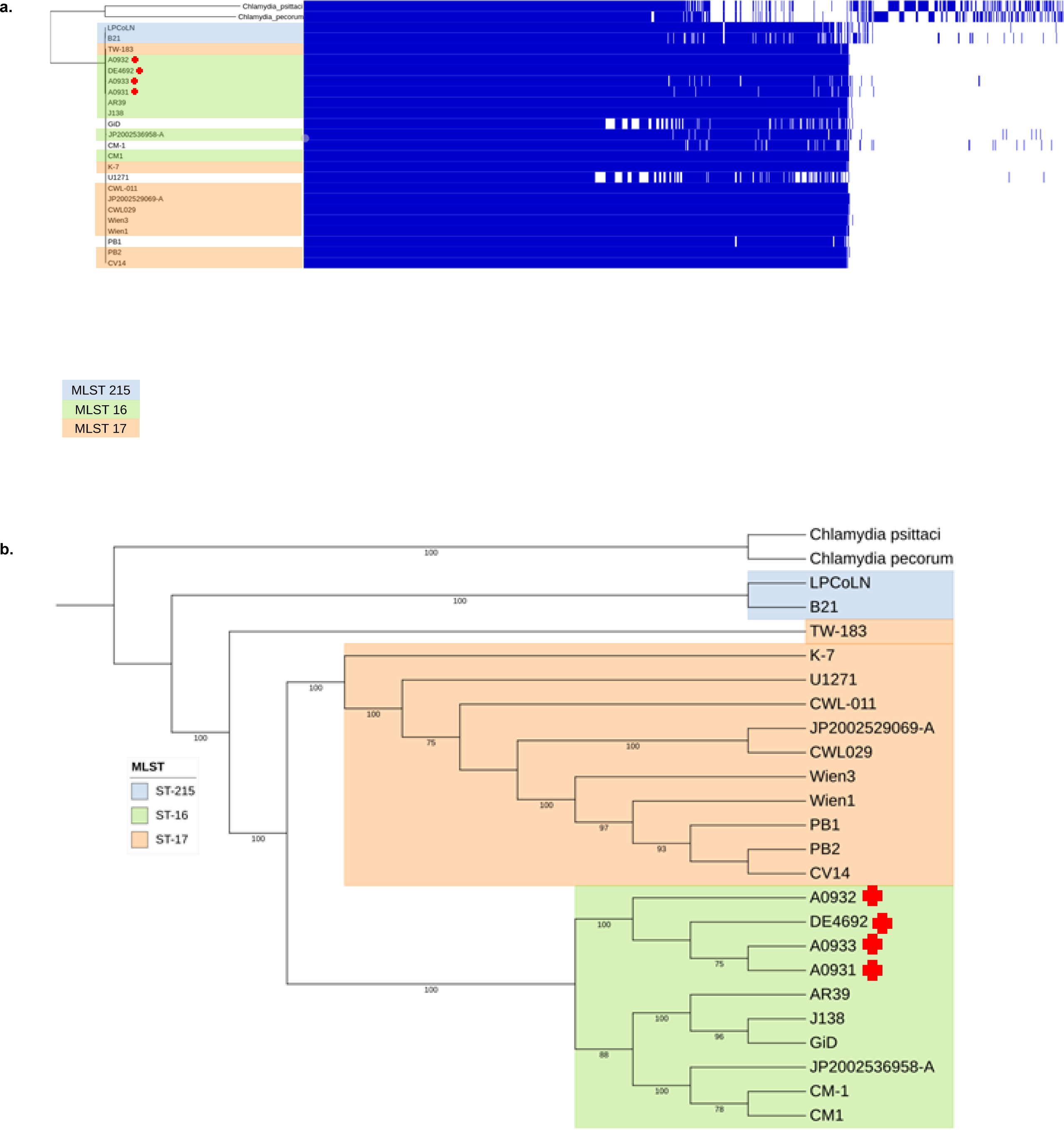
Pangenome analysis showing the cladogram built based on mutations in core genes for *C. pneumoniae* genomes with (a) or without (b) the representation of genes b: genes are represented by blue vertical lines. The four (near) full-length (in 1-12 scaffolds) genomes obtained here are indicated by a red cross.

### APPENDIX, SUPPLEMENTARY TABLES

**Table 1.** Results of the MultiLocus Sequencing Typing (MLST) analysis. The MLST analysis was performed using the PubMLST (https://pubmlst.org/software/bigsdb) web application

| *Chlamydia pneumoniae* genomes | MLST serotype | Markers |  |  |  |  |  |  |
| --- | --- | --- | --- | --- | --- | --- | --- | --- |
|  |  | gatA | oppA | hflX | gidA | enoA | hemN | fumC |
| Reference genomes for the MLST | ST16 | 4 | 7 | 5 | 6 | 5 | 3 | 4 |
|  | ST17 | 4 | 7 | 5 | 7 | 6 | 3 | 4 |
|  | ST18 | 4 | 7 | 5 | 7 | 7 | 3 | 4 |
|  | ST215 | 19 | 19 | 66 | 21 | 17 | 16 | 4 |
| Genomes obtained in the present study |  |  |  |  |  |  |  |  |
| IHUChlampneu_Mars1 (CP172581) | ST16 | 4 | 7 | 5 | 6 | 5 | 3 | 4 |
| IHUChlampneu_Mars2 (CP173416) | ST16 | 4 | 7 | 5 | 6 | 5 | 3 | 4 |
| IHUChlampneu_Mars3 (A0931) | ST16 | 4 | 7 | 5 | 6 | 5 | 3 | 4 |
| IHUChlampneu_Mars4 (A0932) | ST16 | 4 | 7 | 5 | 6 | 5 | 3 | 4 |

